# Effective Implementation of Medicines Shortage Policy: Evidence from Australia’s Serious Scarcity Substitution Instruments

**DOI:** 10.64898/2026.02.02.26345406

**Authors:** Jack Janetzki, Lisa Kalisch-Ellett, Nicole Pratt, Anna Kemp-Casey

**Affiliations:** Adelaide University, (School of Pharmacy and Biomedical Sciences, College of Health), Adelaide, (South Australia), Adelaide

**Keywords:** health policy, drug utilisation, health service administration, pharmacoepidemiology, Therapeutic Goods Administration

## Abstract

**Background:** Medication shortages are a considerable and ongoing issue in healthcare, disrupting consumer access. Since 2021, Australia’s national medicines regulator has issued Serious Scarcity Substitution Instruments (SSSIs), allowing pharmacists to substitute a specific therapeutically equivalent strength and/or formulation of a medicine without prior approval from a prescriber. The impact of SSSIs on utilisation of medicines has not been investigated.

**Objective:** Determine whether SSSIs are effective in addressing medicine shortages and meeting patient need.

**Methods:** This retrospective cohort study used aggregated pharmacy claims to examine the utilisation of 12 medicines which had an SSSI. We calculated the percentage change in defined daily doses dispensed per 1000 population per day in the 11 months after SSSI implementation, compared with the previous two years. A percentage change of less than 20% was used to indicate success.

**Results:** Following product shortages, utilisation fell for 10 of the 12 medicines examined. For eight of these medicines (amoxicillin, cefalexin, estradiol, fluoxetine, insulin degludec with insulin aspart, isosorbide mononitrate, vigabatrin, and warfarin) decreases in utilisation were minimised to <20%. On average, SSSIs where all permitted substitute products were scarce (e.g. abatacept) were associated with larger decreases in use (between -22% and -68%) than those for which none or only some of the substitutes were in shortage (between -45% and +7%, respectively).

**Conclusions:** While product shortages led to decreases in medicines consumption, SSSIs appeared to be successful in limiting decreases. However, SSSIs were less likely to be successful when many of the permitted substitute products were also scarce.

**Key points:**

1. This study is the first to evaluate the effectiveness of Australia’s Serious Scarcity Substitution Instruments (SSSIs) in mitigating medicine shortages using national dispensing data and interrupted time series analysis.
2. Two-thirds of SSSIs successfully limited utilisation declines to less than 20%, with effectiveness strongly linked to the availability of substitute products.
3. By demonstrating variable utilisation outcomes across medicines, this study adds empirical evidence to international debates on substitution policies, suggesting that nationally standardised frameworks like Australia’s SSSIs may function best when supported by robust supply intelligence.
4. SSSIs are a valuable policy tool for maintaining continuity of care during shortages, but timely implementation and ensuring substitute supply are critical for optimal impact.

## 1 Background

Medication shortages are a considerable and ongoing issue in healthcare. In Australia, health professionals and patients are vulnerable to medicine shortages as more than 90% of medicines marketed in Australia are manufactured overseas[1]. The Therapeutic Goods Administration (TGA) is the national medicines regulator and pharmaceutical companies (known as sponsors) are required to report national-level shortages of all prescription medicines and some important non-prescription medicines to the TGA. These shortage notifications are published on the TGA’s Medicine Shortage Reports Database to ensure that health professionals, medicine sponsors and the TGA can take appropriate action to assist patients[2]. When a national medicine shortage occurs, pharmacists may consider several options including, referring the patient to another pharmacy that has stock of the medication, contacting the prescriber for a prescription for an alternative strength of the same product or an alternative medication with the same indication. By law, pharmacists are not able to initiate a change in dosage, strength or quantity of a medication without prior approval from the prescriber. During serious medicine shortages, however, the TGA can authorise a Serious Scarcity Substitution Instrument (SSSI)[3].

An SSSI allows a pharmacist to, for a defined period of time, lawfully substitute a specific therapeutically equivalent strength and/or formulation of a medicine without prior approval from a prescriber or the need for a new prescription if certain conditions are met as outlined in the individual SSSI[3]. For example, on 18^th^ September 2021, a SSSI was made for isosorbide mononitrate due to shortages of the 120 mg tablet[4]. The SSSI was in place until 30^th^ June 2023. During this period pharmacists, when presented with a prescription for isosorbide mononitrate 120 mg tablets, could dispense isosorbide mononitrate 60 mg tablets as an alternative without contacting the prescriber.

SSSIs have been used in Australia since 2021[3], typically when medicine shortages have occurred due to manufacturing issues, supply chain disruption, or sudden changes in demand for medicines. The TGA publish current and lapsed SSSIs on their website[3]. To date, the impact of SSSIs on utilisation of medicines has not been investigated and their effectiveness is unknown. Therefore, the aims of this study were to describe the utilisation trends of selected medicines subject to SSSIs and determine whether they were effective in addressing medicine shortages.

## 2 Methods

### 2.1 Data

We analysed an extract of publicly available Pharmaceutical Benefits Scheme (PBS) data, aggregated by month of supply[5]. The dataset captured dispensings of subsidised prescription medicines and covers all Australian citizens and permanent residents. The data extract included the PBS item number, generic name of the dispensed medicine, strength, formulation, quantity supplied, and month of dispensing. The study period was defined as the period between 24-months prior to the SSSI implementation and the 11 months after, with the month of the SSSI implementation treated as a transition month.

### 2.2 SSSIs examined

Between August 2021 and December 2023, SSSIs were implemented for 12 medicines: abatacept, amoxicillin, cefaclor, cefalexin, estradiol, fluoxetine, insulin degludec with insulin aspart, isosorbide mononitrate, phenoxymethylpenicillin, tocilizumab, vigabatrin, and warfarin. Details of the specific preparations in short supply, and their allowable substitutions, are shown in Table 1. Information regarding date of issue of the SSSI, active SSSI timeframe and permitted substitutions of current and lapsed SSSIs were obtained from the TGA SSSI website[3].

**Table 1:**
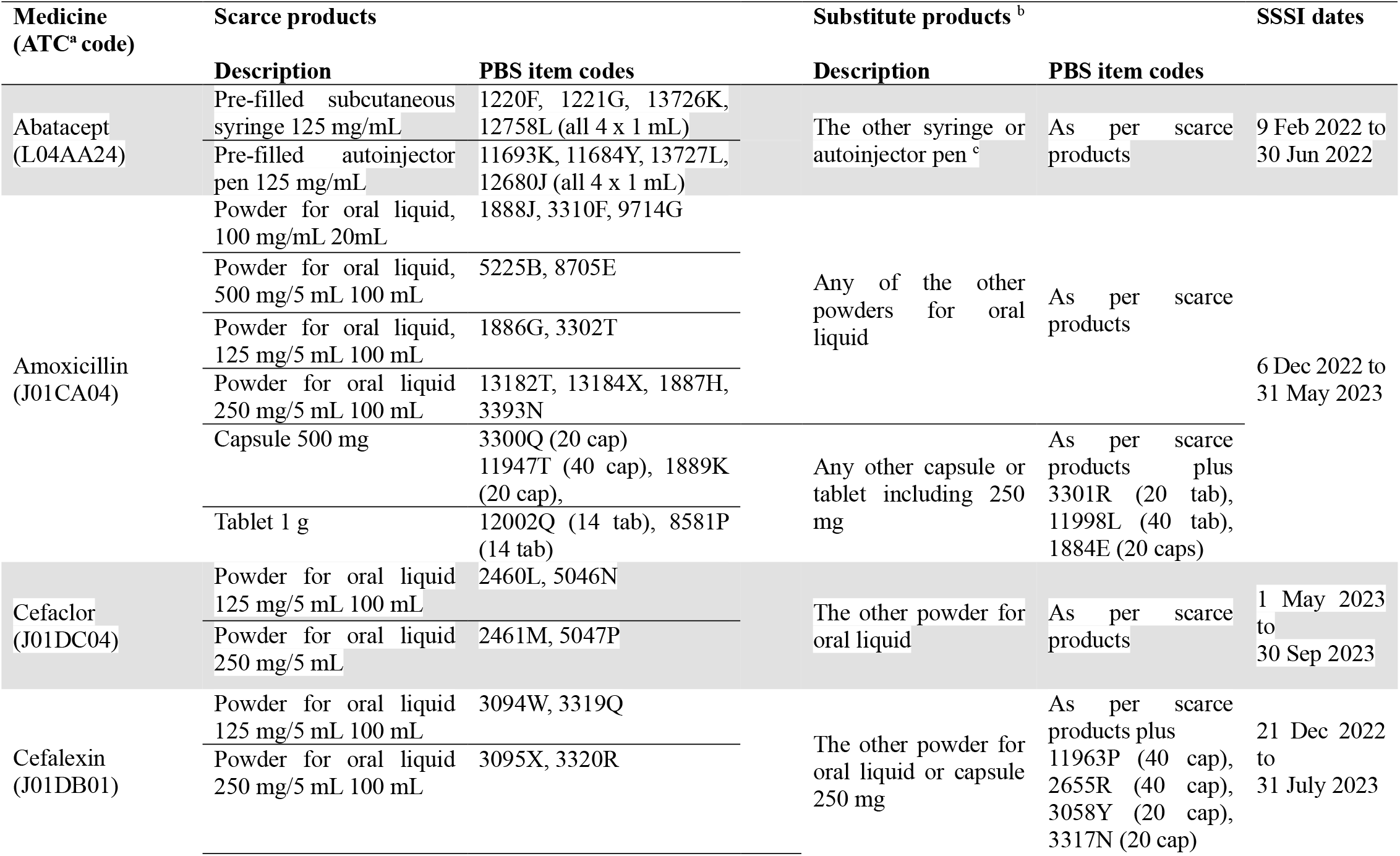
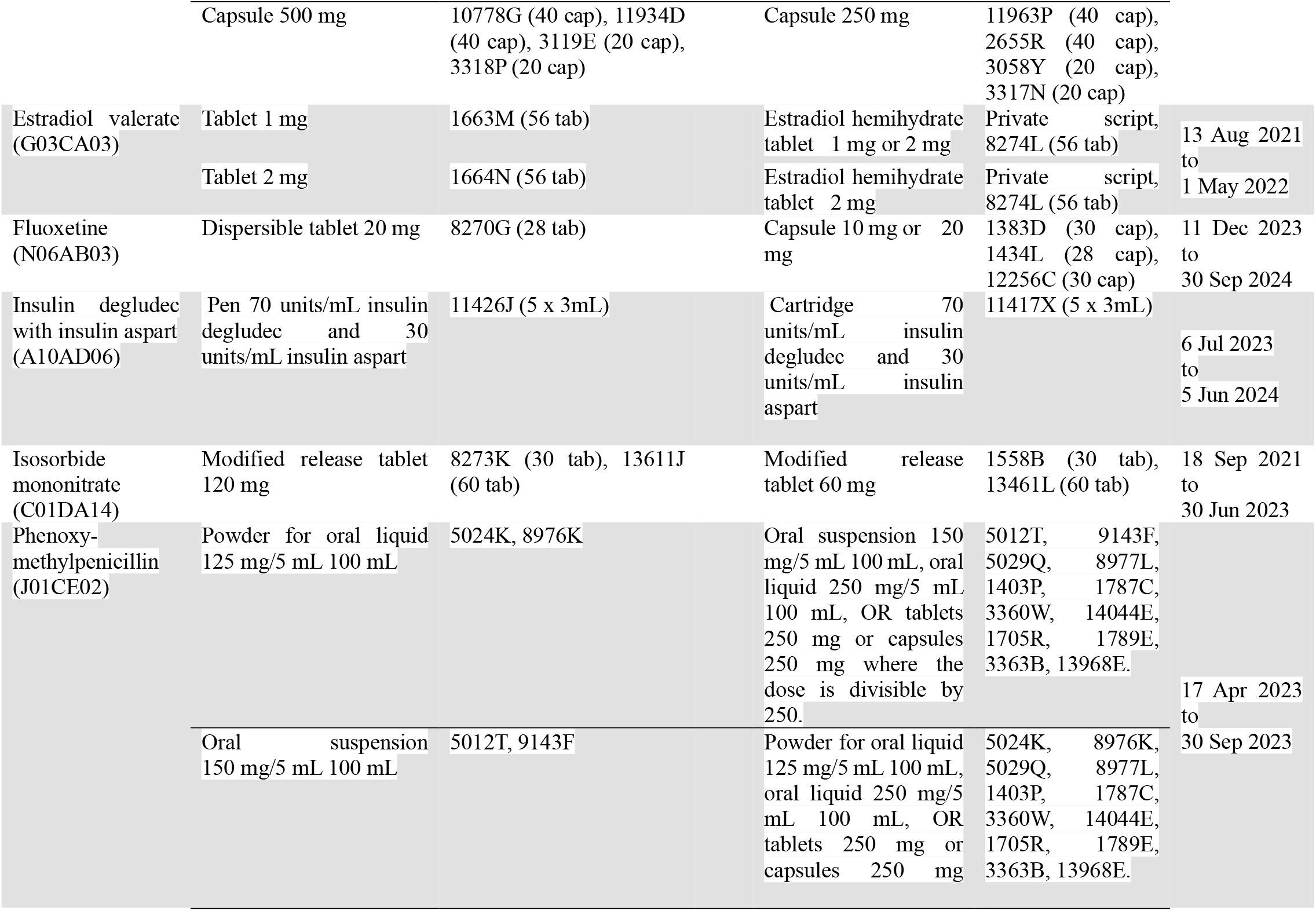
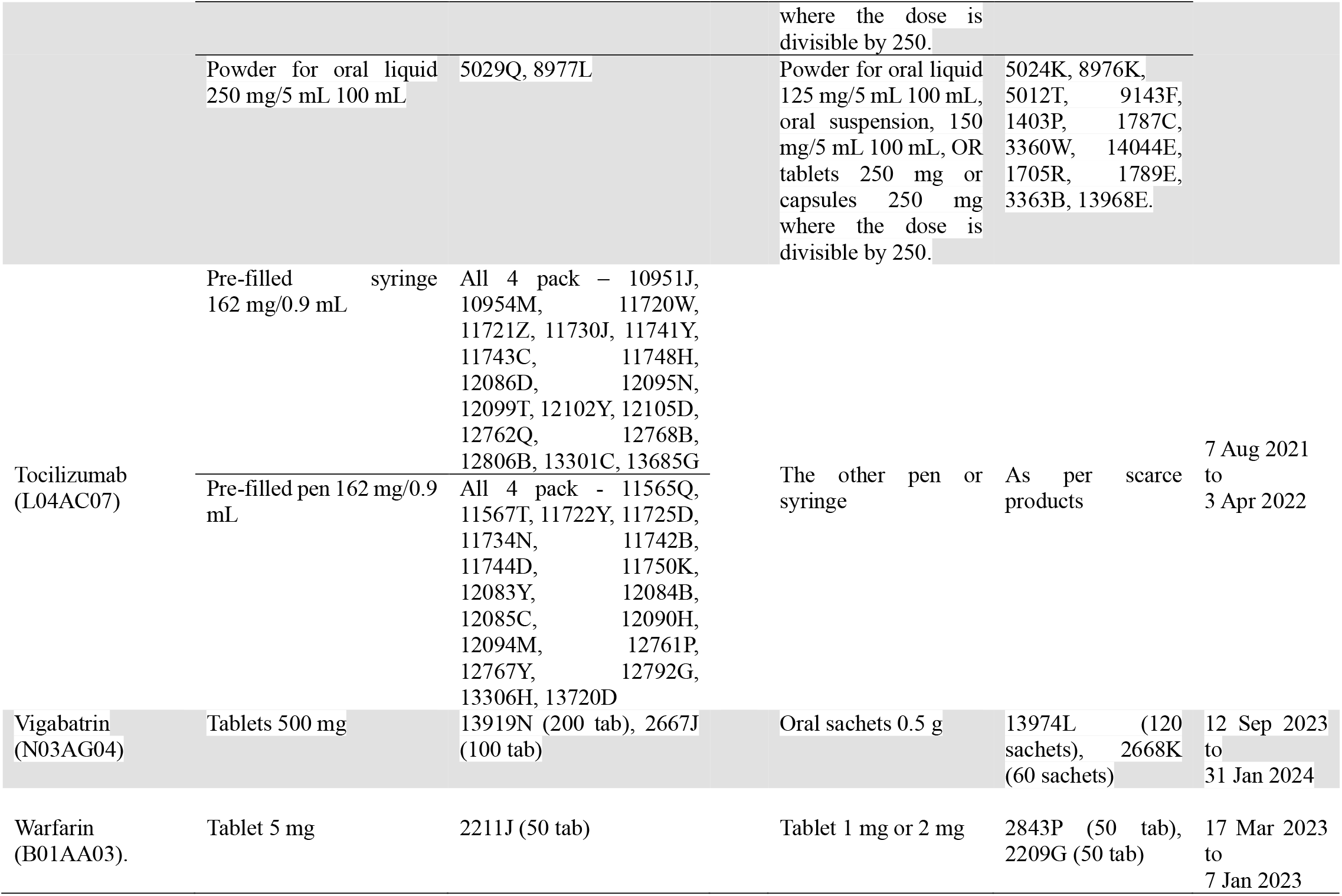
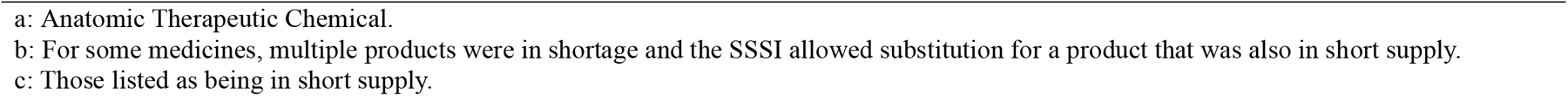
Details of Serious Scarcity Substitution Instruments implemented between January 2021 and December 2023.

## .3 Outcome

The outcome measure for the study was the monthly rate of use of each medicine, expressed as a defined daily dose (DDD) per 1000 population per day (DDD/1000/day). The DDD is an international unit ascribed by the World Health Organization to represent the assumed average daily dose of a medicine required to treat an adult when used for its main indication[6]. Australian population data were obtained from the Australian Bureau of Statistics[7].

### 2.4 Analysis

Interrupted time series analysis was used to examine the monthly rate of utilisation in Australia before and after each SSSI was implemented[8]. We used linear regression models to estimate changes in dispensing in the 24 months before and 11 months after implementation of each SSSI, adjusting for seasonality and autocorrelation. The month of implementation was treated as a transitional month and not included in the models[8]. We calculated the percentage change between the actual dispensing trend and the extrapolated baseline trend for the 11 months after SSSI implementation. The extrapolated baseline trend estimated the dispensings that would have occurred if the medicine shortages had not occurred. We used bootstrapping resampling methods with 5000 iterations to estimate percentile-based 95% CIs for the percentage change[9, 10]. We considered a percentage change in utilisation of less than 20% to indicate success for the SSSI. We based this on the 80% threshold generally used in adherence studies; a convention based on an early study of antihypertensives effectiveness[11]. SAS on Demand for Academics was used for analyses.

### 2.5 Ethics statement

Ethics approval was not required for this study as we utilised publicly available, deidentified aggregated data.

## 3 Results

Australian utilisation of the 12 selected scarce medicines is shown for the two years before and 11 months after implementation of the SSSI (Figure 1). Utilisation of scarce products, substitute products, and the combination of the scarce and substituted products are shown. There are seasonal patterns evident for 7 of the 12 medicines examined. For six medicines (abatacept, fluoxetine, insulin degludec with insulin aspart, isosorbide mononitrate, vigabatrin and warfarin), the seasonal dispensing pattern variation was consistent with the PBS safety net entitlements[12]. Amoxicillin utilisation peaked in the winter months, consistent with use for seasonal illness.

**Fig 1:**
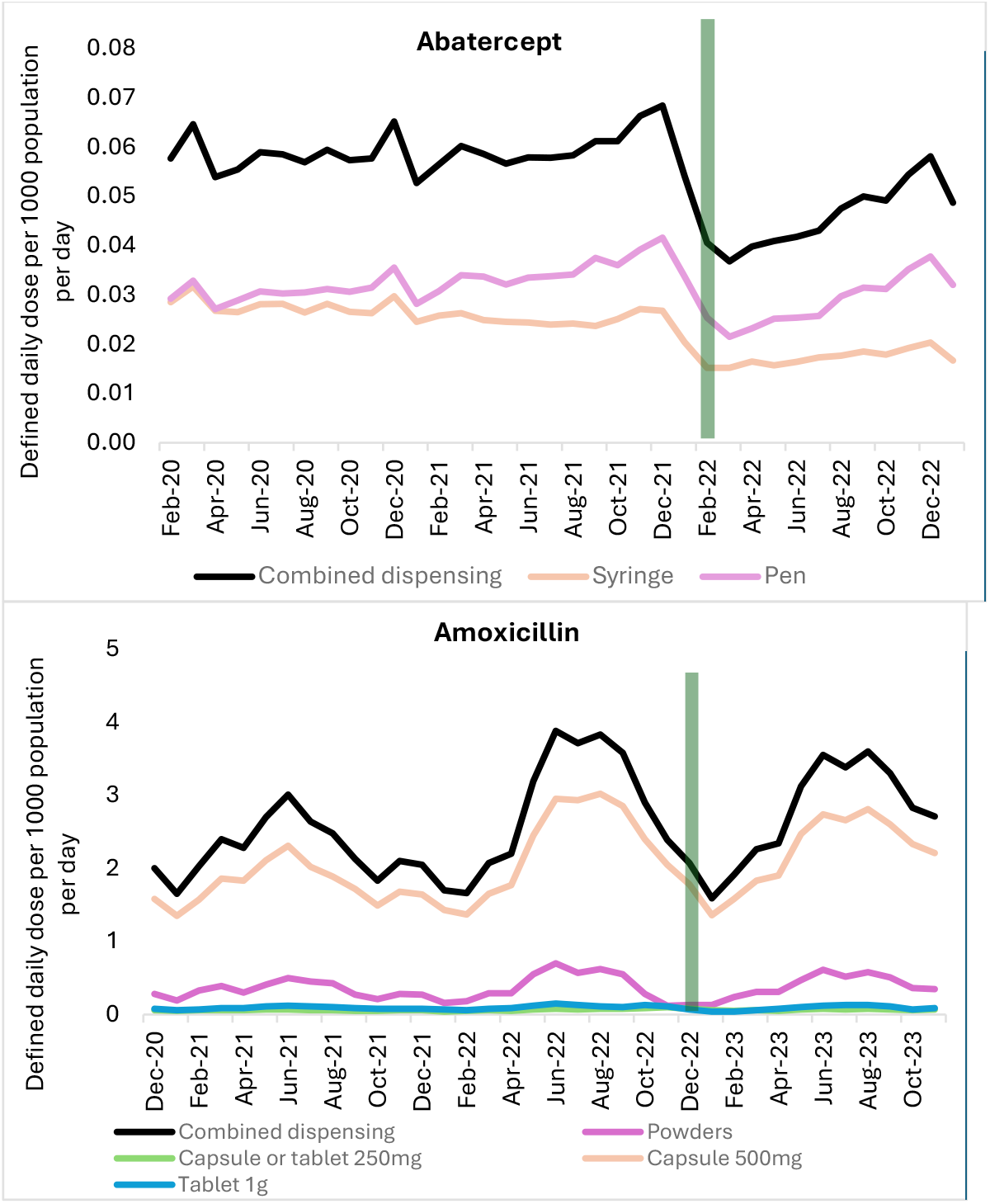
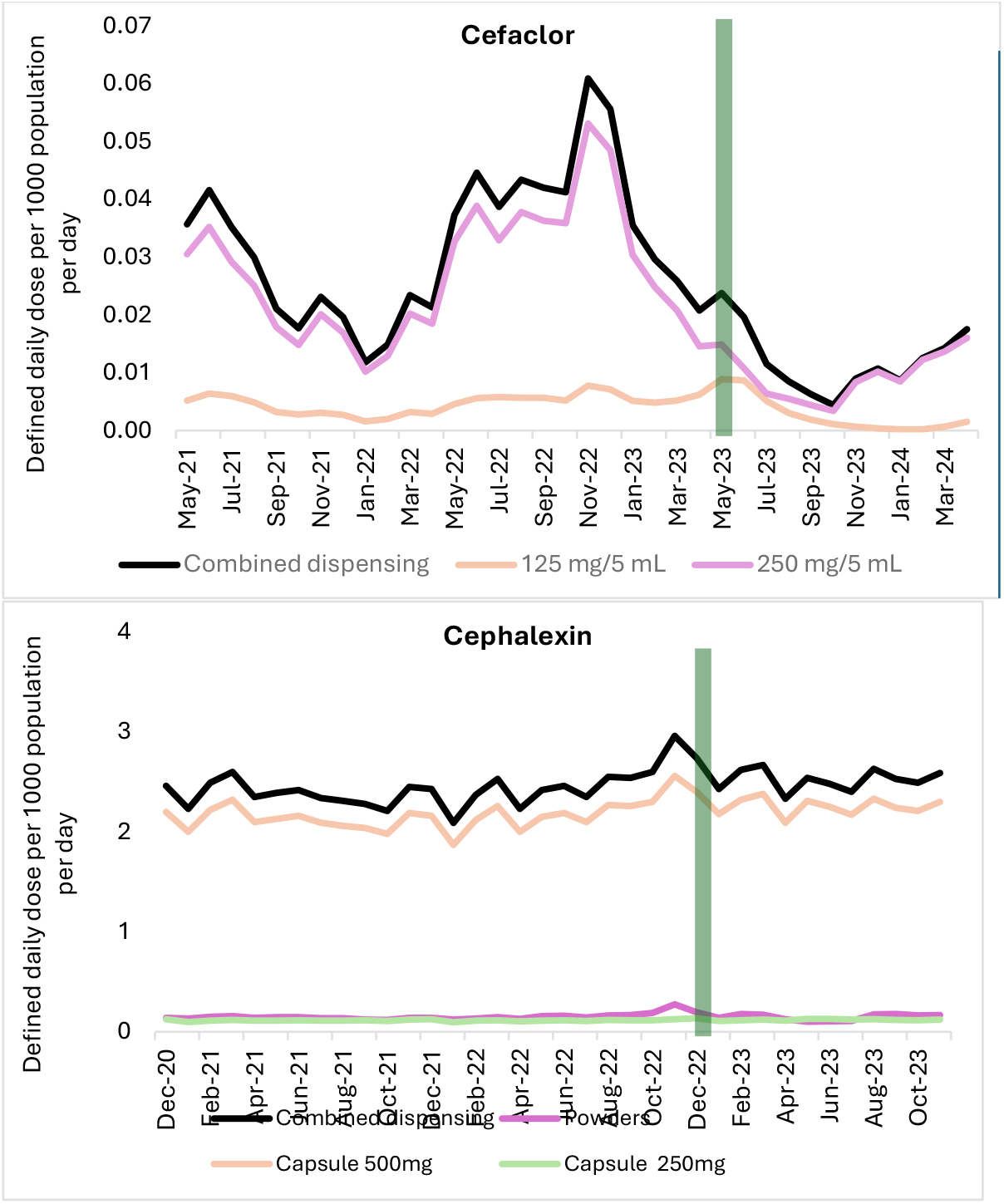
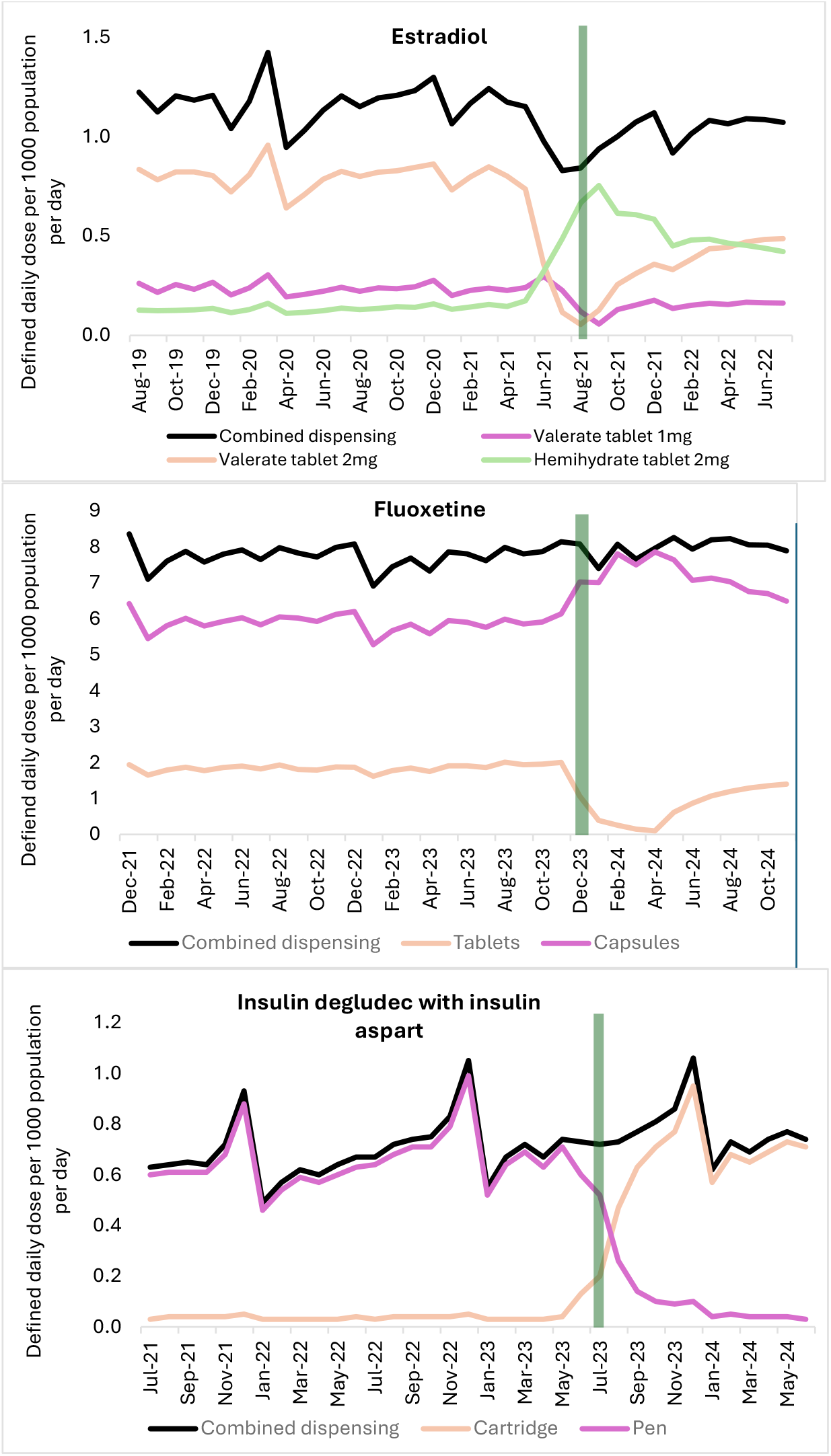
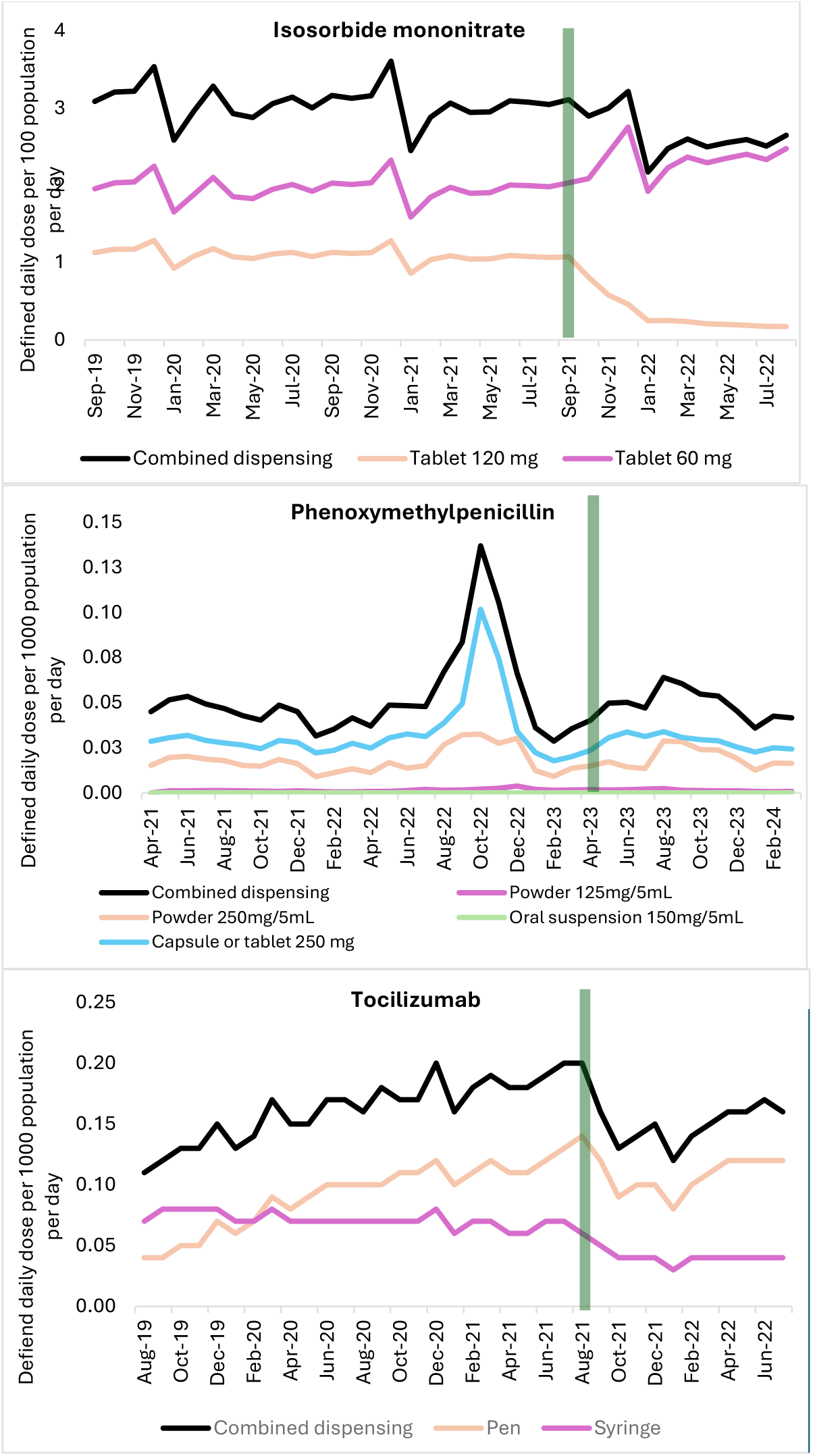
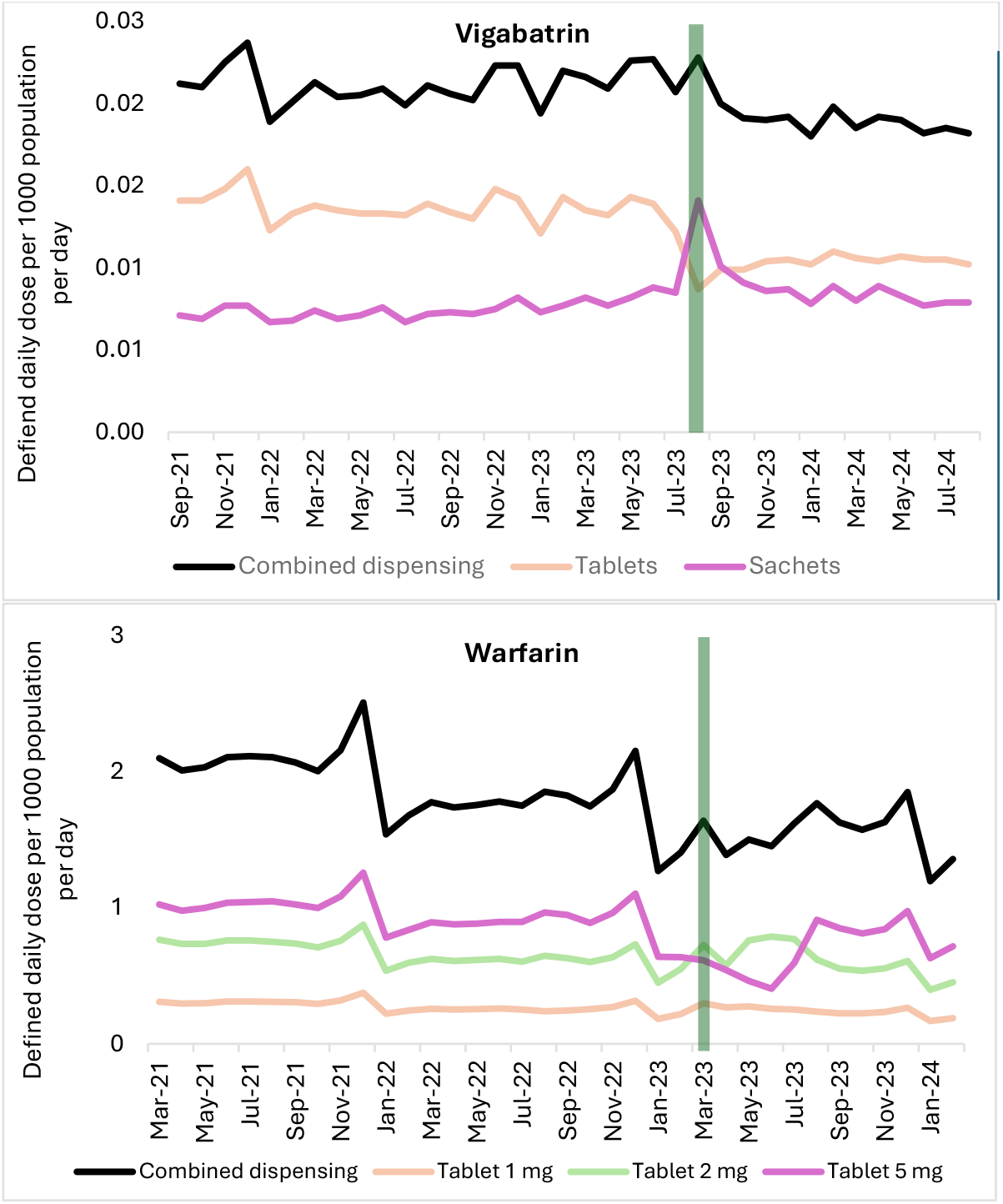
Time series graphs of medicines for which there was a Serious Scarcity Substitution Instrument.

It was observed that cefaclor and phenoxymethylpenicillin had substantial utilisation peaks in October/November 2022 (Figure 1). For six of the 12 medicines, abatacept, estradiol, fluoxetine, insulin degludec with insulin aspart, vigabatrin, and warfarin, decreasing use of the scarce product/s was evident before the SSSIs were implemented.

Figure 2 shows the percentage change in utilisation of the selected medicines for the year after the SSSIs were implemented, compared with the baseline that would have been expected if the shortages had not occurred. Following product shortages, utilisation fell for 10 of the 12 medicines examined. For eight of these medicines (amoxicillin, cefalexin, estradiol, fluoxetine, insulin degludec with insulin aspart, isosorbide mononitrate, vigabatrin, and warfarin) decreases in utilisation were minimised to less than 20%.

**Fig 2:**
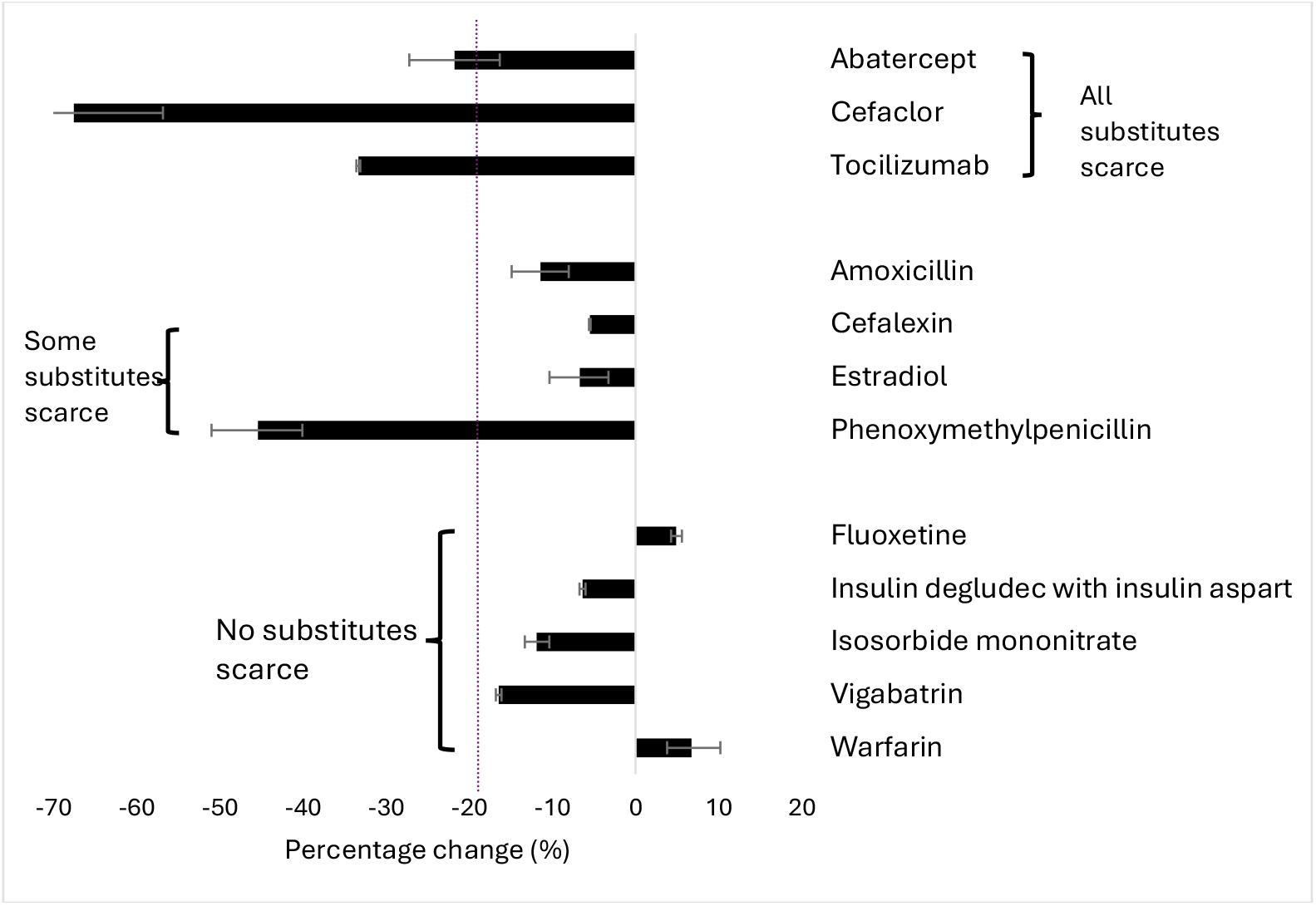
Percentage change (95%CIs) in defined daily doses per 1000 population per day dispensed in the 11 months after implementing the Serious Scarcity Substitution Instrument compared with the previous two years, by medicine. Dotted blue line shows a 20% reduction in use.

The impact on medicine utilisation appeared to be associated with availability of the substitute products. For five of the medicines, fluoxetine, insulin degludec with insulin aspart, isosorbide mononitrate, vigabatrin, and warfarin, none of the substitute products listed in the SSSI were scarce, and the change in use of these medicines was limited to less than 20% (17% (vigabatrin) to an increase of 7% (warfarin)).

When some substitute products were scarce, i.e. amoxicillin, cefalexin, estradiol and phenoxymethylpenicillin, utilisation fell by between 6% and 45% in the year after implementation of the SSSIs (Figure 2). When all substitute products listed in the SSSI for abatacept, cefaclor and tocilizumab were also in scarce supply, utilisation decreased by between 22% and 86% in the year after the SSSIs were implemented.

## 4 Discussion

This is the first study to evaluate the impact of SSSIs on the use of medicines subject to shortages. While the TGA has not specified what ‘successful’ implementation of an SSSI looks like, they are designed to ensure continuity of therapy for patients. Our results indicate that three-quarters of the SSSIs examined were successful in addressing product shortages and potentially maintaining patient adherence.

We identified that availability of substitute products was a key component of SSSI success. When observing duration of shortage there did not appear to be any relationship between duration of shortage and percentage change in use of the scarce medicine or substitutes overall. For many of the medicines (abatacept, amoxicillin, cefaclor, cefalexin, fluoxetine, phenoxymethylpenicillin and vigabatrin), there were multiple shortages listed in the TGA Medicines Shortage Reports Database archive in the two years prior to the individual medicine SSSIs being issued[13]. We noted that for half of the medicines examined the SSSIs appear to have been implemented late (i.e. after product shortages began); however the late implementation of the SSSIs and the number of shortages in the two years prior did not appear to impact on an SSSIs success (analysis not shown).

Medicine shortages are increasingly common and disrupt health care access for consumers[14]. Managing the impact of shortages can also increase the workload on prescribers and pharmacists[15, 16]. Consequently, it is important to understand the effectiveness of programs put into place to manage medicines access in times of scarcity. This is the first study to examine the impact of SSSIs in Australia. Similar schemes are operational in Canada[17, 18], New Zealand[19], and the United Kingdom (UK)[20], although their impacts on utilisation of medicines over time have not been evaluated. Pharmacists in the UK reported finding early scarcity arrangements overly complex and inflexible[21]; however a 12-month Department of Health and Social Care review reported that there were no apparent concerns or negative effects of the policy on the market for prescription medicines[22, 23]. A distinguishing feature of the Australian SSSI framework is its national consistency and legal clarity, with time-limited instruments specifying exact substitute products and conditions of use. This contrasts with more decentralised or discretionary models internationally, which may rely on professional judgement or regional guidance. Our findings suggest that this prescriptive approach may be most effective when substitute products are demonstrably available, highlighting the importance of real-time supply intelligence in policy design

This is the first study to examine the impact of SSSIs on utilisation of medicines with known shortages. This is likely to be of ongoing importance to prescribers, patients and policy makers in the future due to ongoing supply chain issues and international trade instability[14]. Our outcome measure, defined daily dose per 1000 population per day, accounted for underlying changes in the Australian population over time, as well as artefacts that would have occurred had we used prescription counts based on differing pack sizes and doses. Our findings suggest that the effectiveness of SSSIs is contingent not merely on their legal authorisation, but on the underlying availability of substitute products. Future iterations of the SSSI framework may benefit from tighter integration with supply forecasting systems to ensure that authorised substitutes are themselves resilient to shortages.

All studies have limitations. This study used aggregated rather than individual-level data. While SSSIs are intended to support patient need during medicine shortages, our analysis was limited to aggregate utilisation and cannot directly assess patient-level outcomes such as treatment interruption, clinical deterioration, or patient experience. Utilisation therefore represents a proxy indicator of continuity of access rather than a direct measure of patient need being met. The TGA have not specified any medicine utilisation targets for their SSSIs, and despite being based on traditional adherence measures, we acknowledge that our ‘success’ benchmark of <20% is an arbitrary one. We also did not look at substitutions outside of the SSSI which prescribers might have used as substitutes (e.g. other medicines in class)[14]. This would have given insight into prescriber behaviour during shortages but not the success of the SSSIs per se which was our aim.

This study was not designed to estimate the causal effect of SSSIs on medicine utilisation. Rather, we used interrupted time series methods to describe changes in utilisation patterns before and after implementation of SSSIs in the context of known shortages. Without a contemporaneous control group of unaffected medicines, it is not possible to fully account for secular trends or concurrent policy and market factors influencing medicine use. However, by examining multiple medicines across different therapeutic areas and shortage contexts, we were able to identify consistent patterns that provide insight into the conditions under which SSSIs are more or less likely to mitigate utilisation declines. Identifying appropriate comparator medicines unaffected by shortages but subject to similar demand dynamics is challenging in the context of widespread, multi-product supply disruptions occurring during the study period.

## 5 Conclusion

We found that product shortages led to decreases in use for 10 of the 12 medicines we examined. Two-thirds of the SSSIs we examined were successful in limiting utilisation decline to <20%. SSSIs were more likely to be successful when none or only some of the permitted substitute products were also scarce. The findings indicate that SSSIs are an effective strategy for meeting patient medicine needs in times of shortage.

